# The Impact of COVID-19 on Medical Practice: A Nationwide Survey of Dermatologists and Healthcare Providers in Iraq

**DOI:** 10.1101/2020.07.26.20156380

**Authors:** Mohammed Shanshal, Hayder Saad Ahmed, Hayder Asfoor, Raad Ibrahim Salih, Shehab Ahmed Ali, Yusif k. Aldabouni

## Abstract

The COVID-19 pandemic has dramatically changed medical practice worldwide. It posed a significant impact on different health services, including dermatology. A Cross-sectional observational study of 200 healthcare providers and 100 dermatologists (survey 1 and 2, respectively) were conducted to determine the prevalence of occupational skin diseases among healthcare providers working amid the pandemic, and to demonstrate the outbreak’s impact on dermatology practice.

Most healthcare providers (83%) reported hygiene-related hand dermatitis. The rates of PPE- related dermatoses were estimated to be 73%, including pressure injuries (51.9%), acne (33.1%), non-gloves contact dermatitis (29.9%), nonspecific eruption (17.5%), urticaria (9.1%) and skin infections (3.2%). The emerging COVID-19-related cutaneous manifestations were recognized by 20% of surveyed dermatologists, including maculopapular eruption (41.67%), urticaria (37.50%), chilblain (25%) and vasculitis (16.67). Telemedicine was provided by 73% of the dermatologists, and 89% reported minimal use of immunosuppressive drugs amid the pandemic.

---

An ongoing outbreak of coronavirus disease 2019 (COVID-19) caused by SARSCoV-2, started in December 2019. It was first identified in Wuhan, China. The virus then propagated and began to hit other regions around the world until it was declared a global pandemic by the World Health Organization (WHO) on March 11, 2020 ^(1)^.

The emerging COVID-19 virus is phylogenetically linked to viruses that cause severe acute respiratory syndrome (SARS) and Middle East respiratory syndrome (MERS). COVID-19 creates variable degrees of illness, ranging from fever, cough, dyspnea, fatigue, and diarrhea to critical cases of severe acute respiratory distress syndrome ^(2)^.

The COVID-19 pandemic has brought the world to a standstill and placed considerable challenges on healthcare workers, including dermatologists. Telemedicine has gained particular importance, where many of the dermatologic consultations were transitioned into teledermatology services. The current outbreak has led to the ongoing emergence of personal protective equipment (PPE)-related dermatoses and hygiene-related hand dermatitis, particularly among healthcare providers. In addition, due to a shortage of vaccination programs imposed by the COVID-19 lockdown, there is a significant risk for other infectious disease outbreaks, including measles.

## Objectives and methods

A cross-sectional observational study involving two online surveys were conducted for data collection. The purpose of using two questionnaires is twofold:

1. to comprehensively examine the effect of COVID-19 on different aspects of medical practice
2. to concentrate on the virus effect on dermatologic practice.

A 5-item electronic survey was created and sent to 276 randomly selected healthcare providers working in the emergency departments, inpatient wards, and outpatient clinics of the major hospitals in Iraq. The survey assessed the frequency of occupational skin diseases due to hygienic work practices and PPE use among healthcare providers working during the current pandemic. The participants were asked about their gender, age, and the frequency of hygienerelated hand dermatitis and skin complications related to PPE use.

For survey 2, a 12-item electronic questionnaire was sent to 156 randomly selected dermatologists practicing within the COVID-19 era. The dermatologists were asked about the impact of the current outbreak on various aspects of dermatology practice, including the provided medical services, infection control methods, disease epidemiology, and the currently recommended medical and surgical management.

## Results

In the first survey, a total of 200 responses were collected from healthcare providers working amid the COVID-19 pandemic with a response rate of (72.46%). The results of the first survey are summarized in table 1 and 2.

**Table 1:**
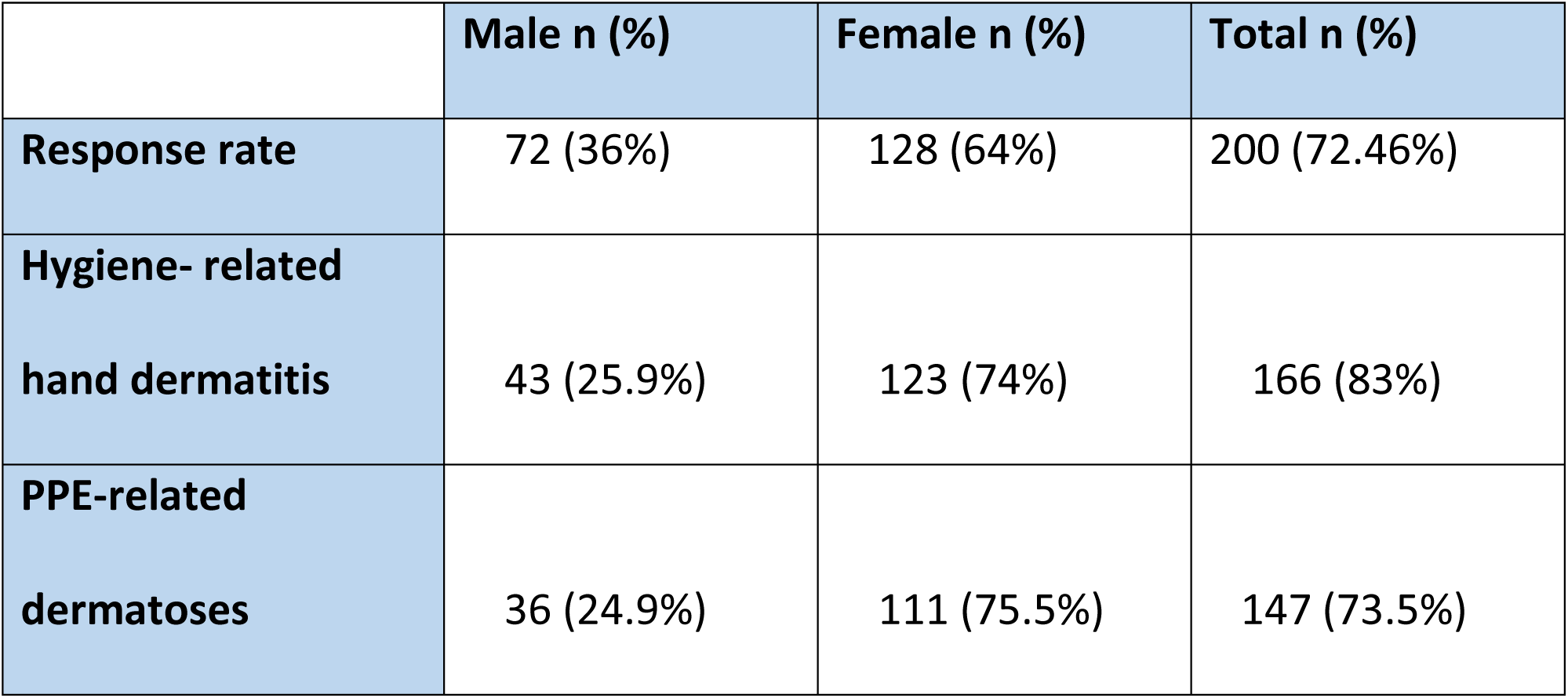
Occupational skin diseases among health care providers working amid COVID-19.

|  | Male n (%) | Female n (%) | Total n (%) |
| --- | --- | --- | --- |
| **Response rate** | 72 (36%) | 128 (64%) | 200 (72.46%) |
| **Hygiene- related**<br>**hand dermatitis** | 43 (25.9%) | 123 (74%) | 166 (83%) |
| **PPE-related**<br>**dermatoses** | 36 (24.9%) | 111 (75.5%) | 147 (73.5%) |

**Table 2:** Types and frequency of PPE-related dermatoses.

|  | Male n (%) | Female n (%) | Total n (%) |
| --- | --- | --- | --- |
| **Pressure injuries** | 14 (17.5) | 66 (82.5%) | 80 (51.9 %) |
| **Acne** | 8 (15.68) | 43 ( 84.3%) | 51 (33.1 %) |
| **Non-gloves CD** | 11 ( 24%) | 35 (76%) | 46 ( 29.9 %) |
| **Non-specific rash** | 5 (18.5%) | 22 (81.5%) | 27 (17.5%) |
| **Urticaria** | 3 ( 21.42%) | 11 (78.57) | 14 (9.1%) |
| **Skin infections** | 3 ( 60%) | 2 ( 40%) | 5 ( 3.2%) |

In the second survey, the questionnaire was completed by 100 practicing dermatologists, providing a 64% response rate. The survey questions and results are illustrated in Tables 3, 4, and 5.

**Table 3.** Impact of COVID-19 on dermatology practice: Survey questions and results.

|  | Yes n= (%) | No n= (%) |
| --- | --- | --- |
| **Do you continue to perform dermatological procedures and biopsies?** | 71 (yes, but at limited capacity)<br>16 ( yes, as before the outbreak) | 13 |
| **Are you still using immunosuppressive to treat skin conditions?** | 68 (yes, but only for limited cases when highly indicated)<br>11 (yes, as before the outbreak) | 21 |
| **Do you diagnose one of the COVID-19 skin manifestations?** | 20 | 80 |
| **Due to COVID-19 related psychological upset, do you notice an increase in relapse rate of some chronic skin diseases?** | 51 | 49 |
| **Do you try to switch immunosuppressive to more conservative options like topical agents/ phototherapy?** | 81 | 19 |
| **Do you get COVID-19 infection during your work?**<br>**(PCR confirmed case only)** | 25 | 75 |
| **Do you provide online/ phone consultations**<br>**(teledermatology)?** | 72 | 28 |
| **Did you work at the respiratory triage or any other COVID-19 related unit?** | 19 | 81 |
| **Did you diagnose a case of measles during the current pandemic?** | 14 | 86 |

**Table 4.**
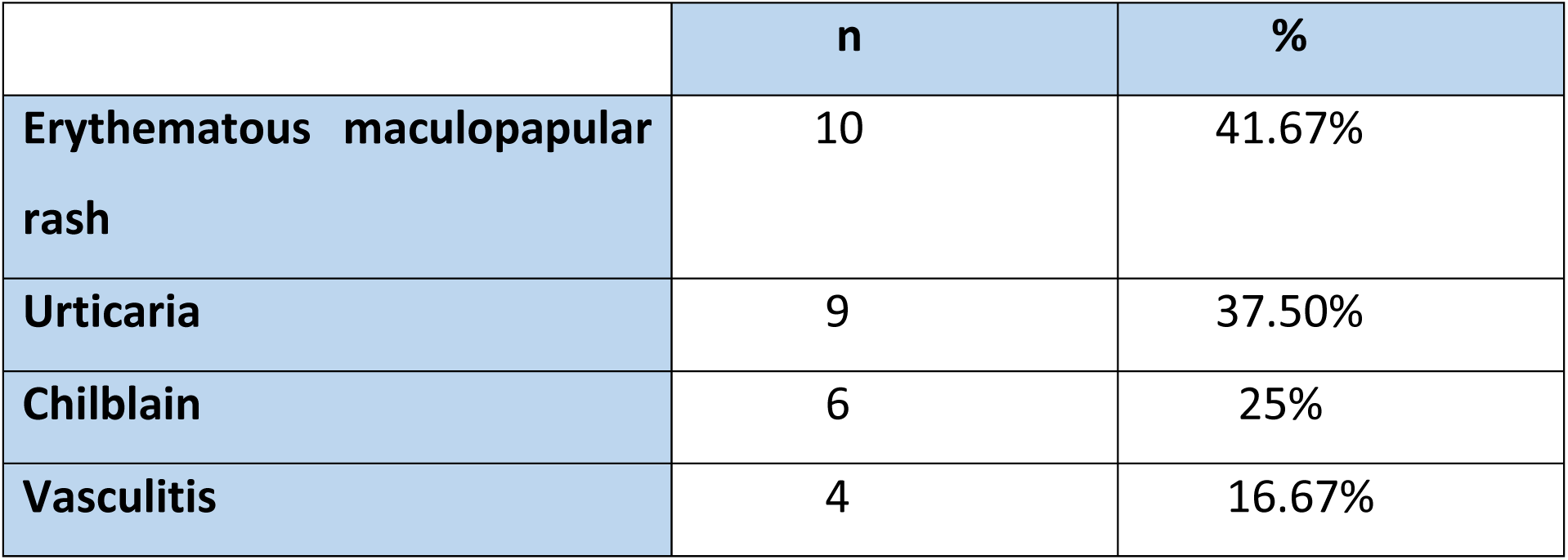
Cutaneous manifestations of COVID-19 diagnosed by dermatologists.

|  | **n** | **%** |
| --- | --- | --- |
| **Erythematous maculopapular rash** | 10 | 41.67% |
| **Urticaria** | 9 | 37.50% |
| **Chilblain** | 6 | 25% |
| **Vasculitis** | 4 | 16.67% |

**Table 5.** Dermatoses aggravated by COVID-19 related psychological stress.

|  | **n** | **%** |
| --- | --- | --- |
| **Psoriasis** | 33 | 64.70% |
| **Atopic dermatitis** | 30 | 58.82% |
| **Rosacea** | 9 | 17.64% |
| **Vitiligo** | 5 | 9.80% |
| **alopecia areata** | 3 | 5.88% |

## Discussion

### Impact of COVID-19 on medical services

We have reviewed the impact of COVID-19 pandemic on the practice of dermatology in Iraq. Although the population of Iraq exceeded 40 million in 2020, based on the latest United Nation data ^(3)^, available statistics indicate that the number of physicians is 7.08 per 10,000 populations which is extremely low when compared to developed countries ^(4)^.

On February 24, 2020, Iraq announced the first confirmed case of SARS-CoV-2 infection. Since then, the spread of COVID-19 has increased. exponentially. By early September, about 260,370 confirmed cases and 7,512 deaths had been reported in Iraq ^(5)^.

The COVID-19 pandemic has targeted every single aspect of medical practice, including dermatology. As Baghdad became the epicenter of the COVID-19 pandemic in mid-March, most dermatology clinics were closed or continued to operate at a minimal capacity. The objective was to provide essential medical care for emergency cases, such as Stevens-Johnson syndrome/toxic epidermal necrolysis, erythroderma, acute angioedema, and lesions worrisome for skin cancer.

With the acute shortage of PPE, medical practitioners clenched their teeth with each patient, wondering if they had been exposed to the virus already. COVID-19 patients may present with a wide range of dermatologic manifestations with or without typical respiratory signs ^(6) (7)^.

Of the 100 dermatologists involved in the survey, 20% reported diagnosing one or more COVID-19 related skin manifestations. These have included the most commonly diagnosed diseases: erythematous maculopapular eruption (41.67%), urticaria (37.50%), chilblains (25%), and vasculitis (16.67%) (Figure 1).

**Figure 1:**
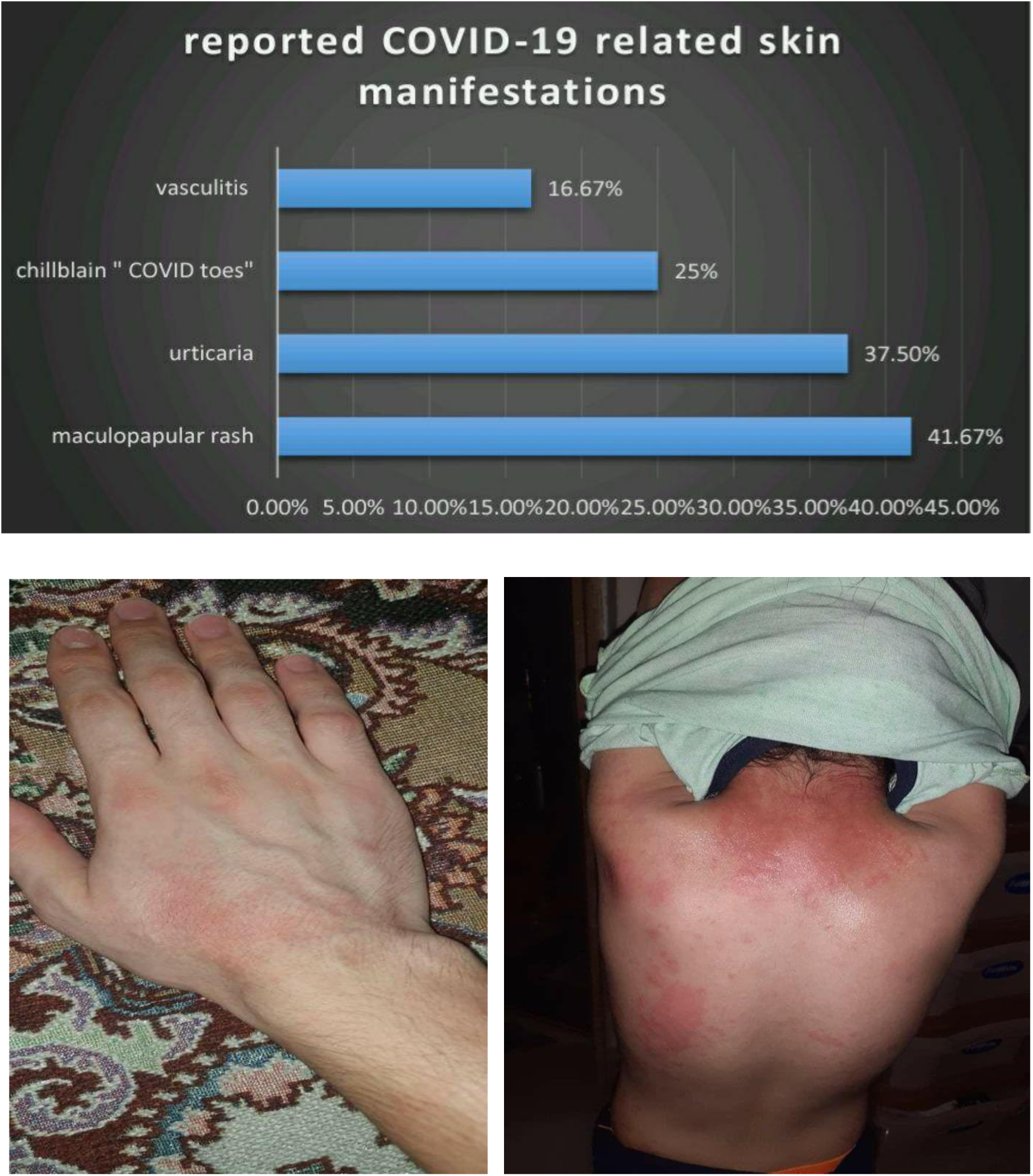
COVID-19 related cutaneous manifestations. **(A)** The approximate frequency of diagnosed COVID-19 related skin manifestations reported by dermatologists. **(B)** 25 years old male with erythematous maculopapular eruption involving the extremities **(C)** 12 years old girl with generalized urticarial eruption.

### Impact of COVID-19 on infection control

The dermatology team tried to reduce face-to-face consultations in both public hospitals and private practice clinics, limiting them to emergency patients. Nonemergent presentations, including melasma, acne, vitiligo, and stable psoriasis, were diverted to telemedicine platforms to avoid the unwarranted risk of spreading infection. Despite these measures, about 25 dermatologists contracted COVID-19 during routine patient care.

The survey revealed that during the outbreak, 72 % of practicing dermatologists provided teledermatology consultations through telephone and/or online social network applications. Because dermatology diagnosis depends primarily on visual characteristics, dermatologists had already been leaders in providing telemedicine services even before the crisis, leading to greater success with this experience.

In order to reduce the risk of infection spread while attending dermatology clinics, several measures have been implemented including stressing commitment to social distancing and wearing a mask, the pre-examination measurement of the patient’s temperature, and exclusion of the respiratory symptoms at the hospital entrance respiratory triage unit, in addition to the meticulous disinfection of equipment^(8)^(Figure2).

**Figure 2:**
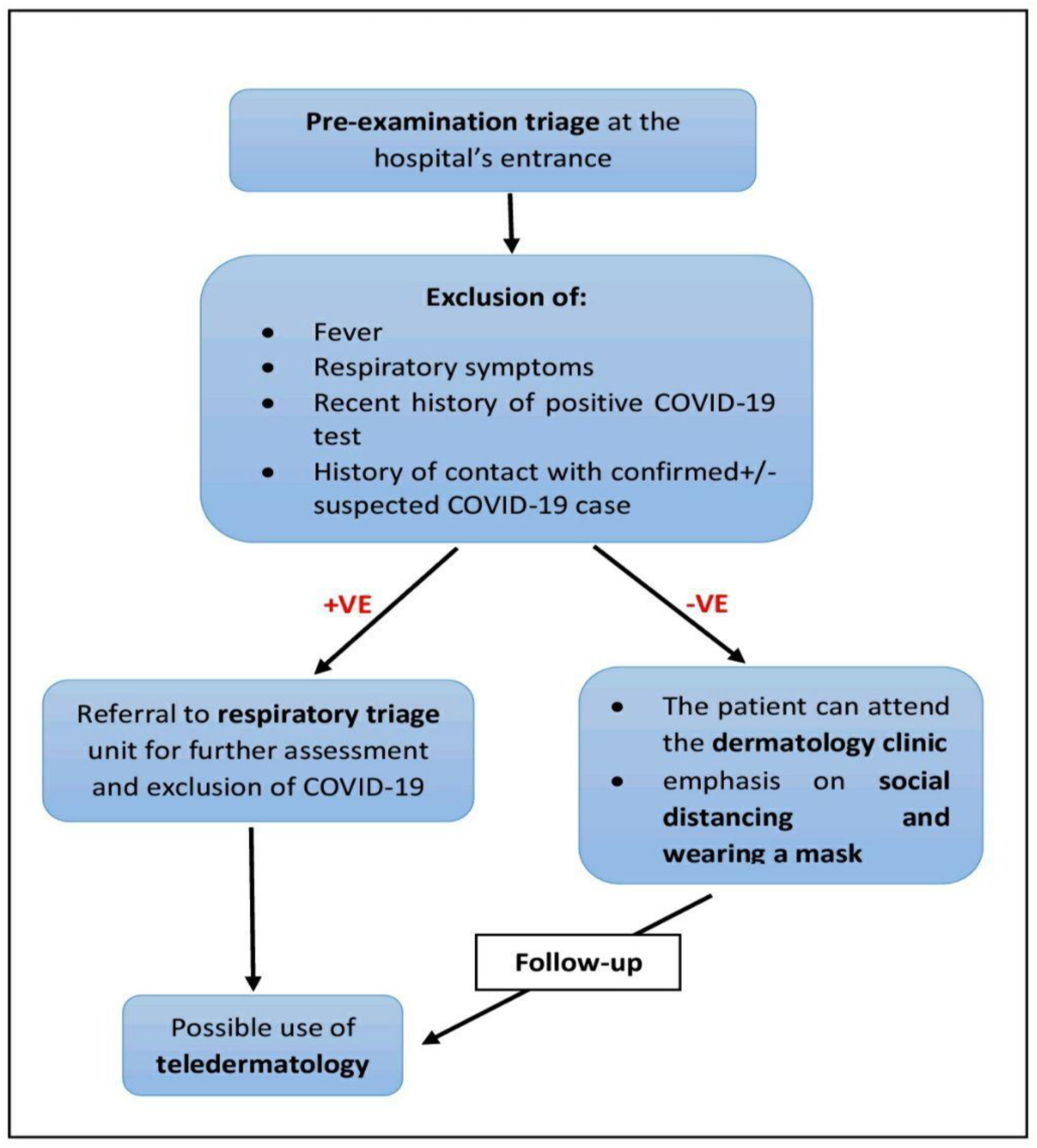
COVID-19 pre-examination triage steps. Flowchart represents the pre-examination triage to decrease the risk of infection spread while attending the dermatology clinic and utilizing teledermatology for initial +/-follow-up assessment.

### Impact of COVID-19 on skin disease epidemiology

#### ➢ Hand dermatitis

Due to frequent handwashing, prolonged use of gloves, and overuse of disinfectants, the incidence of hand dermatitis among healthcare staff and the general population has dramatically increased (Figure 3).

**Figure 3:**
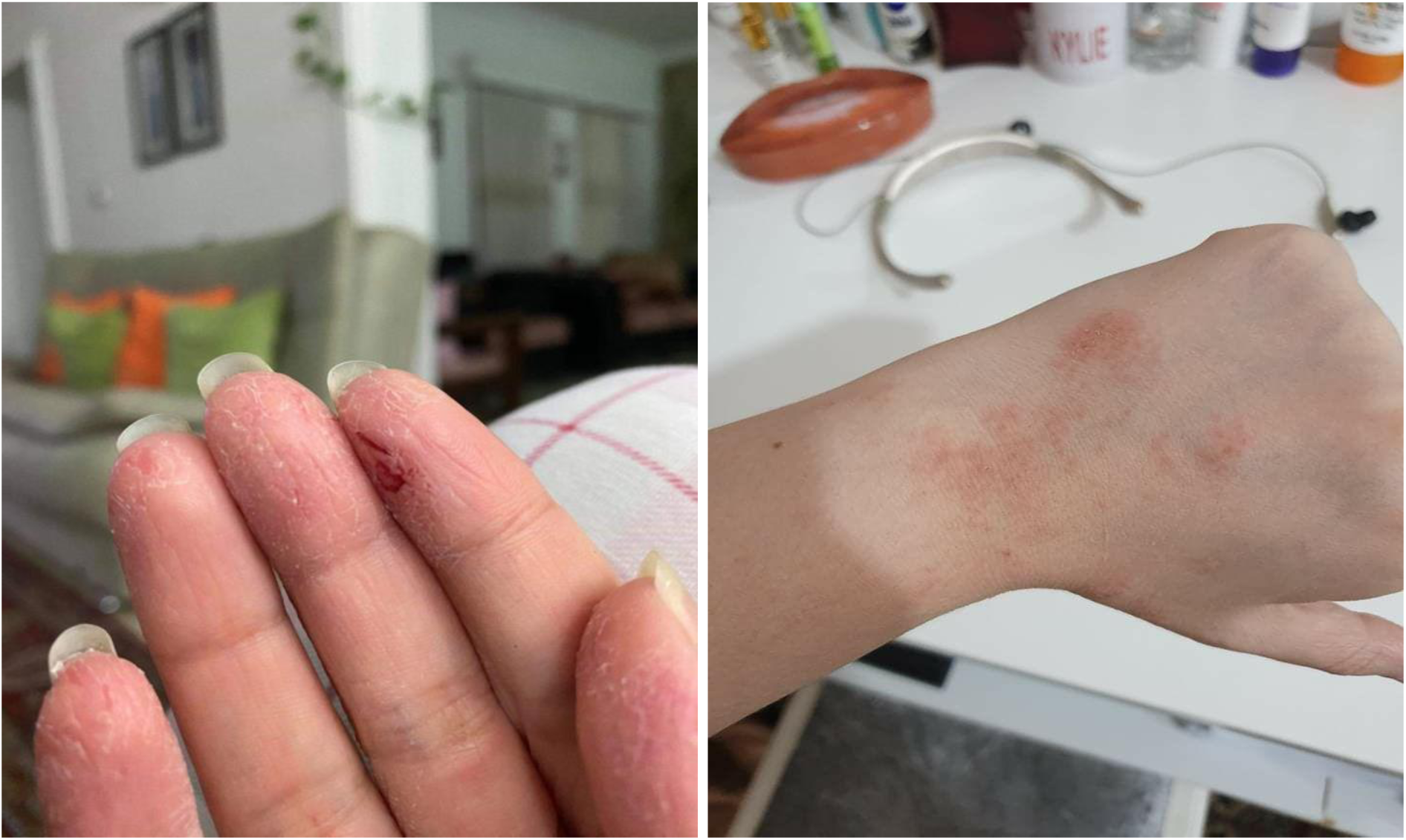
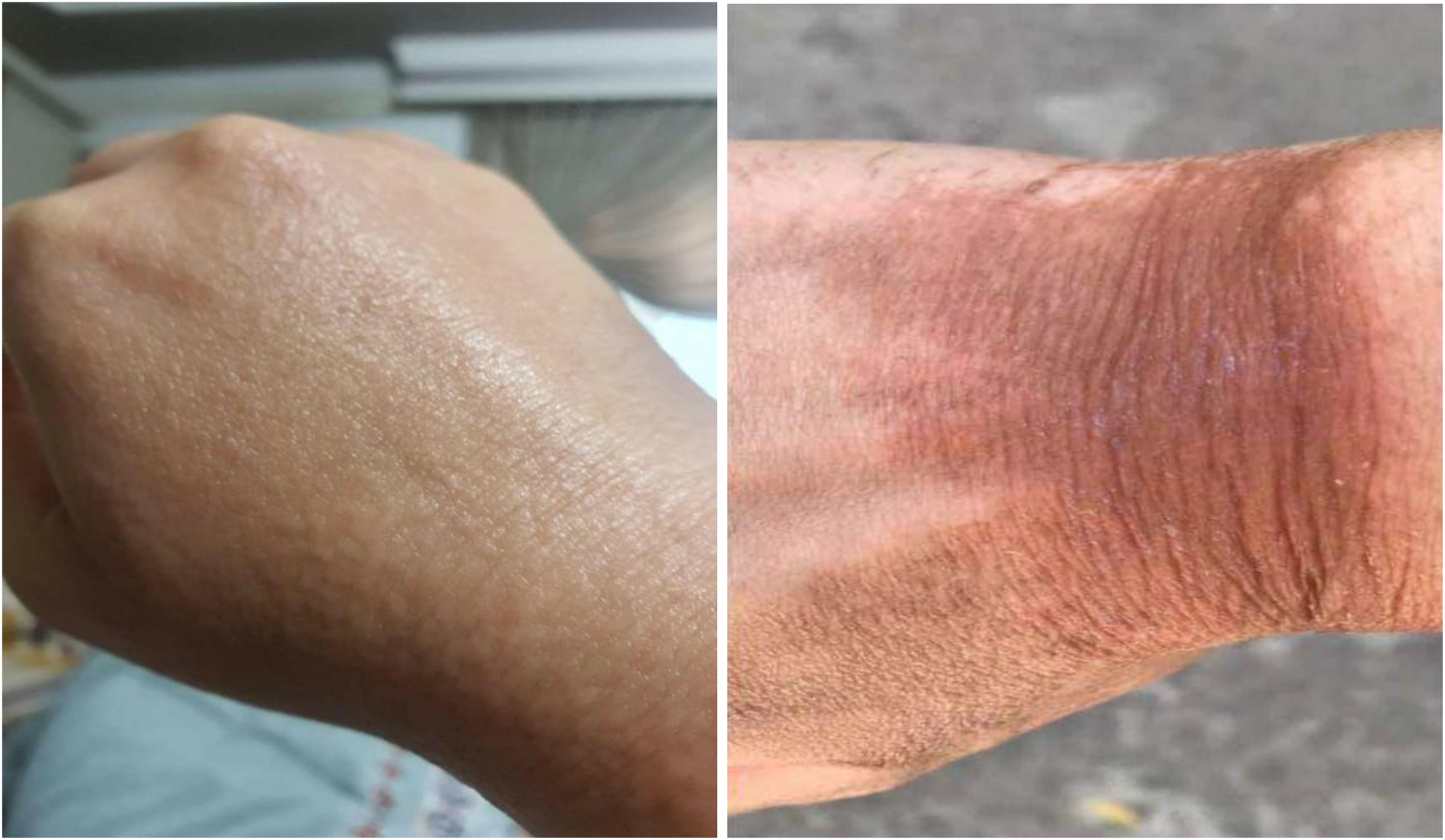
Hygiene-related hand dermatitis. **(A) (B)** Eczematous contact dermatitis **(C)** Papular contact dermatitis **(D)** Post-inflammatory hyperpigmentation following severe disinfectants-related dermatitis

In this survey, hand dermatitis was reported in 83% of healthcare providers working during the current pandemic. Another study reported a comparable result of gloves-related hand dermatitis (88.5%) among healthcare providers working in Hubei Province hospitals amid the outbreak ^(9)^. A pre-outbreak epidemiologic study in mainland China found the frequency of hand dermatitis to range from 14.3% to 23.8% across hospital departments, far from those recorded during the COVID-19 outbreak ^(10)^.

Several measures can be used to decrease the incidence of hand dermatitis, including the use of mild soaps (syndets) that are equally effective in preventing the viral spread and frequent use of moisturizers and physical barriers, especially after handwashing to protect the skin and decrease irritation. Depending on the severity of the lesions, established hand dermatitis should be treated with emollients and/or topical steroids ^(11).^

#### Personal protective equipment (PPE)-related dermatoses

Prolonged contact with PPE such as masks, protective gowns, face shields, and goggles has led to an unprecedented emergence of PPE-related dermatoses among healthcare providers working amidst the current pandemic. PPE-induced skin injuries are attributed to mechanical friction, long-term occlusion, maceration, and allergic contact reactions ^**(12)**^.

Among the surveyed healthcare providers, 147 (73.5%) reported PPE-related dermatoses. Of these, 111 (75.5%) occurred in women and 36 (24.9%) in men. The median age was 31 years (range 24-47 years; standard deviation 4.94 years). The frequency of PPE-related dermatosis reported in this survey includes: pressure injuries (51.9%), acne (33.1%), non-gloves contact dermatitis (29.9%), nonspecific dermatitis and itching (17.5%), urticaria (9.1%), and skin infections (3.2%) (Figure 4). Folliculitis can occur at the site of skin friction, and the inflamed follicles may become infected with bacteria, especially *Staphylococcus aureus*. Friction caused by surgical masks can also trigger reactivation of the *Herpes simplex* virus.

**Figure 4:**
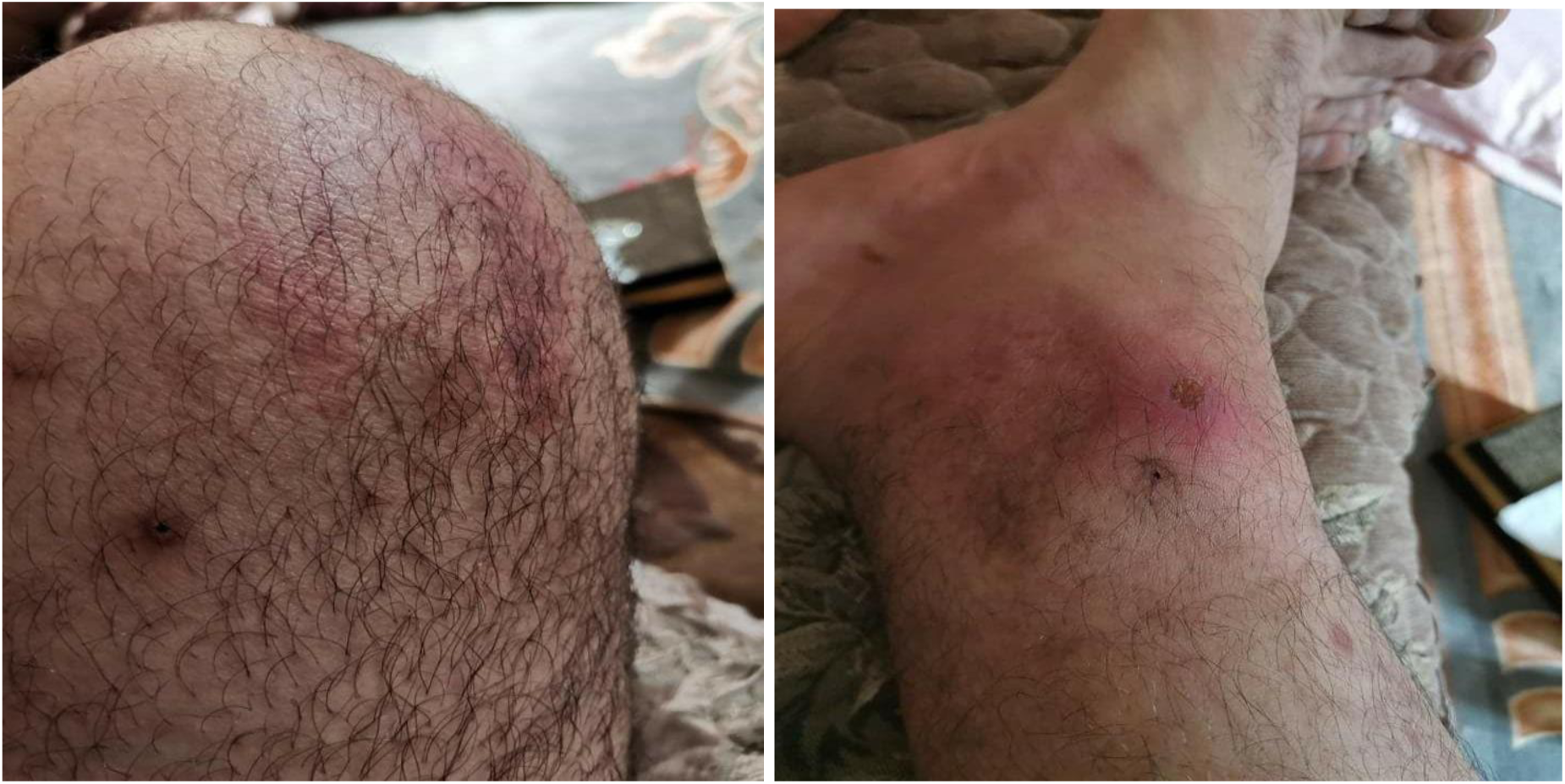
Spectrum of PPE-related dermatosis among healthcare provides working in COVID-19 era. **(A)** Pressure marks and frictional acne **(B)** Mask –induced contact dermatitis (**C)** and **(D)** Folliculitis and herpes simplex reactivation triggered by surgical mask friction **(E)** and **(F)** gown-related contact dermatitis and urticaria **(G)** Pressure injuries and nonspecific rash **(H)** mask-related frictional acne **(I)** Goggles-related nasal bridge dermatitis and hyperpigmentation.

The frequency of PPE-related cutaneous complications among healthcare workers in the Hubei Province during the COVID-19 era included adverse skin reactions associated with the prolonged wearing of PPE, including N95 mask, latex glove, and protective clothing, were 95.1%, 88.5% and 60.7%, respectively ^(8)^.

During the SARS outbreak, the prevalence of PPE-related dermatosis among Singapore health care workers included the findings that 35.5% of the staff who used masks regularly developed acne (59.6%), facial itching (51.4%), and dermatitis (35.8%), while 21.4% who used gloves regularly developed adverse skin reactions, including dry skin (73.4%), itching (56.3%), and dermatitis (37.5%) ^(13)^.

In an analysis of incident case reports from UK dermatologists, non-glove PPE related dermatoses between 1993 and 2013 revealed that 15.5% of the affected patients were health and social workers ^(14)^.

General measures to reduce skin damage associated with the PPE include wearing suitable PPE, avoiding prolonged use wherever possible, and working in a cooler environment. In the areas of maximum tension, dressings can be applied to the skin to redistribute pressure and reduce friction. Frequent use of moisturizers is advisable to protect and repair the skin barrier, and adequate cleansing is necessary to prevent secondary infections ^(15)^.

Mild pressure injuries with intact epidermis usually heal spontaneously without intervention. Small bullae can be kept intact, while larger ones can be aspirated to ease the associated pressure. Eczematous lesions are treated with moisturizers and/or topical steroids, depending on the severity of the lesions. Antihistamines may be used for associated pruritus. For resistant cases, a patch test may be necessary to identify the allergen involved. Mask-related frictional acne can be treated with topical benzoyl peroxide, retinoids, and antimicrobials, similar to acne vulgaris ^(16)^.

#### ➢ Chronic skin disease relapse

It is currently enigmatic how COVID-19 can affect those with chronic skin conditions such as psoriasis and how it can affect these individuals’ treatment. The psychologic impact of the crisis, patients’ reluctance to attend a clinic appointment, limited use of immunosuppressive drugs, and shortage of drugs lead to a perceptible increase in the rate of relapse and exacerbation of several chronic skin diseases ^(17)^. Relapse rates of psoriasis, atopic dermatitis, rosacea, vitiligo, and alopecia areata were noticeably increased as observed by 64.70%, 58.82%, 17.64% 9.80%, and 5.88% of dermatologists, respectively.

Patients should be strongly advised against stopping their medications without consulting their physicians ^(18)^. The importance of supporting patients with chronic dermatologic conditions during the current crisis cannot be overemphasized.

#### ➢ Preventable infectious diseases

The WHO expressed great concern about the spread of previously controllable infectious diseases like measles. Derangement of immunization services caused by the ongoing COVID-19 pandemic is likely to further to decrease measles vaccination coverage by an additional 20%, leaving the most susceptible children at risk for highly infectious disease outbreak ^(19)^. About 14% of the surveyed dermatologists recorded new cases of measles amid the current pandemic; hence, health authorities are called to emphasize the importance of identifying and reaching children with missed vaccine doses to avoid an inevitable outbreak.

### Impact of COVID-19 on dermatologic treatments

Several expert opinions have recommended restricting the use of immunosuppressive agents during the current pandemic ^(20) (21)^. Most dermatologists in the survey reported reduced use of non-biologic immunosuppressants and limited their prescription to selected cases, particularly when alternative options are exhausted, and only after COVID19 symptoms have been excluded. In this condition, the dose should be kept to the lowest acceptable level that still adequately controls the disease.

Due to the uncertainty of whether biologics may place patients at a higher risk of COVID-19 or more dramatic disease course, the initiation of biologic therapy in the current crisis is unadvisable. Alternative therapeutic approaches, particularly in vulnerable patients, may be considered ^(22)^. Patients already receiving scheduled biological therapy, who do not exhibit respiratory symptoms or test positive for COVID-19 may continue their treatment while underscoring the importance of COVID-19 preventive measures, like social distancing and selfquarantining ^(23)^.

### Impact of COVID-19 on surgical dermatology

Most of the non-essential dermatological procedures and biopsies were postponed to minimize the risk of spreading COVID-19 infection. Skin surgery was limited to a few undeferrable biopsies and high-risk skin cancer removal when the risk of delay exceeded the risk of exposure to the virus (Figure 5). Some hospitals have applied specific internal protocols to protect their patients and staff, including testing patients for COVID-19 before their scheduled surgeries and procedures, providing full PPE to all operating staff, and solicitous intraoperative environment sterilization and waste disposal ^(24)^.

**Figure 5:**
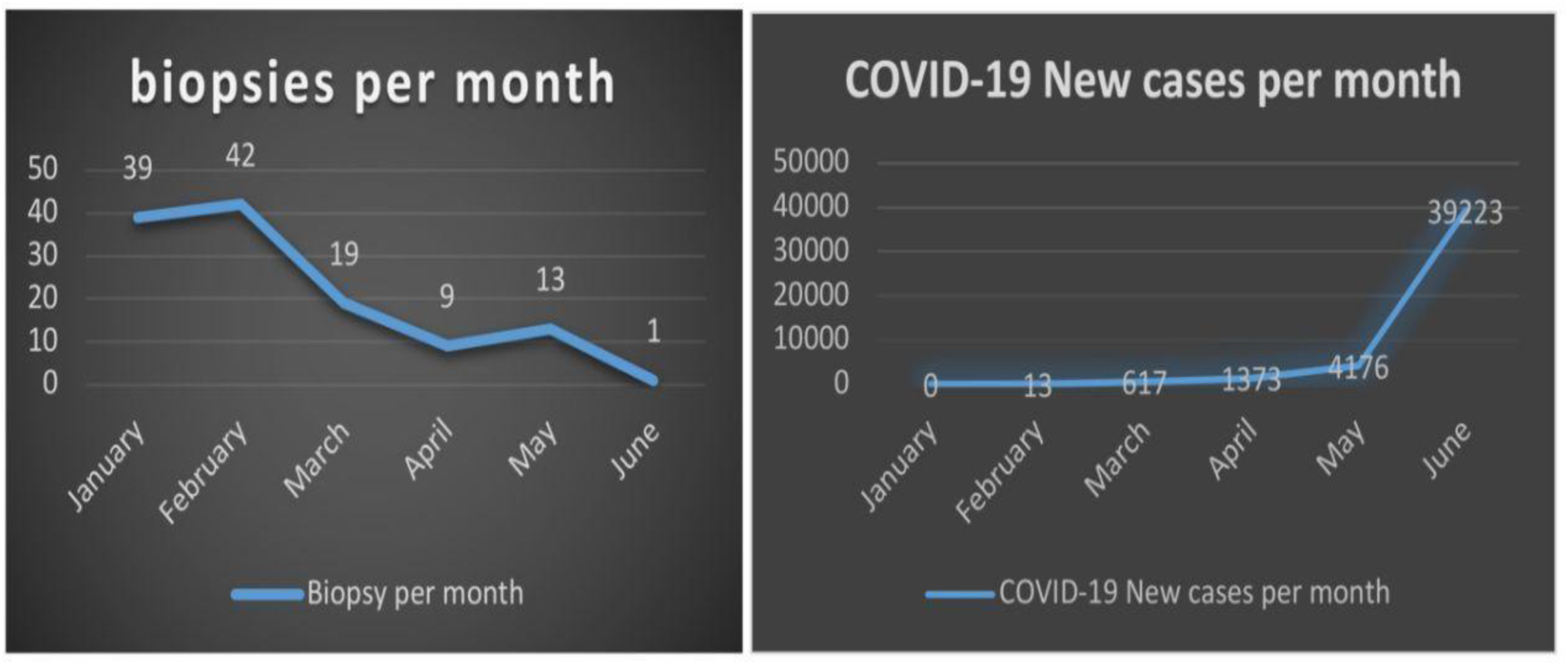
COVID-19 and surgical dermatology. A dramatic decrease in the number of skin biopsies at Baghdad Teaching Hospital in response to the increasing numbers of COVID-19 patients.

### Working on frontlines

Baghdad has been hit particularly hard by COVID-19, where almost half of the confirmed cases were recorded. The dermatology staff felt the call of duty and were actively involved in the respiratory triage at the emergency department, where they continued to help patients and support their fellow physicians. Nineteen percent of the dermatologists surveyed reported working in the respiratory triage or other COVID-19 related units.

## Conclusion

The COVID-19 related preventive measures are associated with an upsurge in occupational skin diseases related to excessive handwashing, overuse of disinfectants, and prolonged utilization of personal protective equipment. The pandemic’s psychological impact, the fear of attending hospitals, and the shortage of medications have led to an increase in the relapse rate of common chronic skin diseases like psoriasis and atopic dermatitis. The dermatologic management plans were overshadowed by the current public health crisis, where dermatologists continued to collaborate with their physician fellows in the battle against COVID-19 infection.

## Data Availability

The used surveys are available for readers.

https://drive.google.com/drive/folders/1eH6inpIkkWpkJVmHYVNqmxXaBXQo9bJN?usp=sharing

## Acknowledgement

Dr. Jhan Adwer Darzi edited our presentation, and Dr. Mohammad Fawzi Al Sultan assisted in the data collection.

